# Remote Cognitive-Motor Training Combining Mental and Physical Practice for Freezing of Gait in Parkinson’s Disease: a Randomized Controlled Trial

**DOI:** 10.64898/2026.05.07.26352678

**Authors:** Paloma Rodrigues da Silva, Karina Yumi Tashima Honda, Larissa Bitarães Rodrigues dos Santos, Julya Morais Garcia, Bruna Hiromi Tateyama da Silva, Luiza de Mattos Aranha, Maria Elisa Pimentel Piemonte

## Abstract

**BACKGROUND:** Freezing of gait (FOG) is a disabling feature of Parkinson’s disease (PD). Although physical practice (PP) improves gait, maintaining gains remains challenging. Mental practice (MP), including Dynamic Neuro-Cognitive Imagery (DNI), may enhance gait control, but evidence on remote combined interventions is limited.

**PURPOSE:** To investigate whether adding MP grounded in DNI principles to remote physical practice supports greater and more sustained improvements than remote physical practice alone in people with PD and FOG.

**METHODS:** A prospective, single-blind, parallel-group randomized controlled trial was conducted. Forty-three participants with idiopathic PD and FOG were randomized to an experimental group (EG, n = 20) or control group (CG, n = 23), stratified by cognitive performance. Both groups received 10 remote sessions over 6 weeks. All performed structured physical practice targeting gait components; the EG additionally performed MP based on DNI, while the CG performed time-matched seated stretching. Assessments were conducted at baseline (BI), post-intervention (AI), and 30-day follow-up (FU). The primary outcome was Rapid Turns Test performance; secondary outcomes included FOG severity, motor aspects of daily living, mobility-related quality of life, and global cognition.

**RESULTS:** All randomized participants were included in intention-to-treat analyses; 38 completed all assessments. Significant group × time interactions were found for Rapid Turns Test duration (p = 0.0019) and FOG time (p = 0.0108). Both groups improved short-term, but only the EG maintained gains at follow-up. Additional interactions favored the EG for mobility-related quality of life (p = 0.001) and global cognition (p = 0.0018). Self-reported FOG improved over time in both groups (p < 0.001) without between-group differences, while motor aspects of daily living showed a time effect only (p = 0.001).

**CONCLUSION:** MP based on DNI principles may enhance retention of gains when combined with remote physical practice, supporting its use as an adjunct in FOG rehabilitation.

**Trial registration:** This trial is registered at ClinicalTrials.gov with trial registration number NCT06957405 (registered on April 25, 2025).

**Protocol and statistical analysis plan:** The full trial protocol and statistical analysis plan are available upon request from the corresponding author.

**Data sharing:** The datasets generated, used and analyzed during the trial are or will be available from the corresponding author upon reasonable request.

**Funding and conflicts of interest:** This article was produced as part of the activities of FAPESP Research, Innovation and Dissemination Center for Neuromathematics (grant #2013/07699-0, Sao Paulo Research Foundation). Co-author PRS received individual support from FAPESP (grant number 2025/14403-7). The authors declare no conflict of interest.

## Introduction

### Background and rationale / Objectives

Freezing of gait (FOG) is one of the most disabling manifestations of Parkinson’s disease (PD), substantially compromising mobility, functional independence, and quality of life [1,2]. People with FOG show greater postural instability, increased risk of falls, and more severe limitations in activities of daily living, making FOG a major contributor to disability in PD [3,4]. According to the recent definition proposed by the International Freezing of Gait Consensus Group (ICFOG), FOG is characterized by paroxysmal episodes in which individuals are unable to step effectively despite the intention to walk, and may occur during gait initiation, turning, walking, or transitions across different locomotor contexts [5].

Although clinically relevant, the pathophysiology of FOG remains incompletely understood. Current evidence indicates that FOG is multifactorial and should not be interpreted solely as a motor impairment, but rather as the result of dysfunctional interactions among motor, cognitive, limbic, cortical, subcortical, and brainstem networks [6,7]. A central component of this process is the decline in automatic gait control, related to impaired basal ganglia-mediated motor regulation and disrupted interaction with brainstem locomotor centers [7]. As automaticity deteriorates, people with PD increasingly rely on attentional and executive resources to regulate gait, shifting from automatic to more goal-directed control [8]. This compensatory strategy may support walking under simple conditions, but becomes vulnerable during turning, dual-task walking, environmental complexity, or emotional stress, when cognitive demands exceed available resources and FOG episodes may be triggered [9,10].

Neuroimaging studies support this interpretation by showing that people with PD and FOG often recruit compensatory cortical networks during locomotor and lower-limb motor tasks, particularly involving prefrontal, premotor, supplementary motor, parietal, and associative regions [11,12]. These areas are involved in motor planning, attentional allocation, inhibition, visuospatial processing, and executive regulation. However, compensatory recruitment in people with FOG appears to be unstable or inefficient, suggesting that rehabilitation strategies capable of strengthening internal motor representations and top-down control may be clinically relevant [7,11].

Mental practice, based on motor imagery, may represent one such strategy. Motor imagery refers to the mental simulation of an action without overt execution, whereas mental practice involves repeated and structured rehearsal of imagined movements, usually incorporating visual and kinesthetic components [13]. Importantly, mental practice engages neural systems that overlap with those activated during physical practice, including premotor, supplementary motor, parietal, and associative cortical regions [14,15]. These regions are also implicated in compensatory mechanisms used by people with PD to overcome the loss of gait automaticity [7,11]. Therefore, mental practice may help reinforce internally generated motor plans, improve anticipation of movement demands, and support cognitive-motor strategies when automatic gait control is compromised [14,16].

This rationale is particularly relevant for FOG because freezing episodes often occur when internally generated locomotor commands fail or when adaptation to contextual demands is required. Mental practice may allow individuals to rehearse gait-related strategies in challenging situations without the immediate risks associated with physical execution. In people with PD and FOG, imagined gait tasks have been associated with recruitment of visuospatial and executive regions, suggesting that imagery may engage alternative neural pathways involved in motor planning and compensatory control [17]. Dynamic Neuro-Cognitive Imagery extends conventional mental practice by emphasizing embodied, kinesthetic, anatomical, spatial, and affective aspects of movement representation, potentially increasing the vividness and functional relevance of imagined actions [16,18]. Previous studies suggest that Dynamic Neuro-Cognitive Imagery may improve imagery ability, motor performance, cognition, and body schema in people with PD and in non-clinical populations [19,20].

However, mental practice is unlikely to replace physical training. Instead, its effects appear to be more clinically meaningful when combined with physical practice [21]. While mental practice may support internal simulation, motor planning, and anticipatory control, physical practice provides sensorimotor experience, task repetition, feedback, and adaptation to real movement constraints. The combination of mental practice and physical practice may therefore act through complementary mechanisms: mental practice reinforcing internal representations and cognitive-motor strategies, and physical practice consolidating these strategies through overt execution [14,22]. This combined approach may be especially relevant for FOG, in which successful walking depends not only on producing steps, but also on anticipating triggers, reorganizing posture, adapting to environmental demands, and recovering from gait interruption.

In parallel, telerehabilitation has emerged as a promising strategy to improve access to long-term rehabilitation for people with PD. Access to specialized care is often limited by mobility restrictions, transportation difficulties, geographic distance, financial constraints, and the need for repeated sessions over time [23]. Remote delivery may help overcome these barriers by allowing supervised practice in the home environment, where real-life mobility challenges occur. In addition, home-based telerehabilitation may facilitate repeated, context-specific practice, support adherence, and promote transfer of trained strategies to daily activities [24,25,26]. For interventions based on mental practice and physical practice, remote delivery may be particularly suitable because imagined and physical rehearsal can be performed safely with limited equipment and adapted to the participant’s own environment.

Despite these advances, important gaps remain. Evidence on mental practice-based interventions in PD is still heterogeneous, with substantial variability in protocols, outcomes, and target populations. Few studies have specifically investigated people with FOG as the primary target population, and even fewer have combined mental practice grounded in Dynamic Neuro-Cognitive Imagery principles with structured physical practice. Moreover, the effects of this combined cognitive-motor approach delivered remotely remain insufficiently explored.

Therefore, the primary aim of this study was to investigate the effects of a remote intervention combining mental practice, grounded in Dynamic Neuro-Cognitive Imagery principles, with physical practice, compared with physical practice alone, on FOG severity in people with PD. Secondary aims were to examine the effects of the intervention on motor aspects of daily living, mobility-related quality of life, and global cognitive performance. We hypothesized that both interventions would improve FOG severity, but that the combined mental practice and physical practice intervention would produce greater and more sustained benefits by reinforcing internal motor representations and supporting top-down control processes involved in gait regulation.

## Methods

### Patient and public involvement

Patients and the public were not involved in the design, conduct, reporting, or dissemination plans of this research.

### Trial Design

A prospective, single-blinded, parallel-group, randomized clinical trial conducted in agreement with CONSORT guidelines for developing randomized trials. The study was approved by the Research Ethics Committee under protocol number 7.253.458. All procedures will comply with the ethical standards of the 1964 Helsinki Declaration and its later amendments, as well as with the Brazilian National Health Council Resolution 466/2012.

### Changes to trial protocol

No important changes were made to the protocol after trial commencement.

### Trial setting

Participants were consecutively recruited through contacts from the AMPARO network (www.amparo.numec.prp.usp.br) using a non-probability sampling method. Clinical information was obtained from healthcare system records of the services where participants received medical care for PD. To ensure that the information accurately reflected participants’ current clinical status, only records dated within the previous six months were considered. All clinical data were subsequently verified with participants and, when applicable, their family members. Details of dopaminergic treatment, including medication type and daily dosage, were recorded to calculate the levodopa equivalent daily dose (LEDD). The study was conducted remotely, and both assessments and interventions were delivered via video-call platforms, performed between May and December of 2025.

## Eligibility criteria

### Eligibility criteria for participants

Participants were eligible if they met the following criteria: clinical diagnosis of idiopathic PD according to the UK Parkinson’s Disease Society Brain Bank criteria [27]; current use of dopaminergic medication; presence of FOG, defined by a positive response to item 1 of the New Freezing of Gait Questionnaire (NFOG-Q) [28]; ability to walk independently at home; access to the internet and to a device capable of video calls; and willingness to participate in the study.

Participants were excluded if they had other neurological disorders; severe cardiovascular and/or respiratory conditions; uncorrected visual or auditory impairments; unstable medical or psychiatric conditions that could interfere with participation or outcomes; changes in antiparkinsonian medication within 30 days before enrolment; or participation in concurrent physiotherapy specifically targeting gait or FOG. Participants were also excluded if they had cognitive impairment likely to compromise comprehension of verbal instructions, defined as a Telephone Montreal Cognitive Assessment (T-MoCA) score below 12 [29,30], or insufficient motor imagery ability, defined as a Kinesthetic and Visual Imagery Questionnaire-20 (KVIQ-20) score of 20 or lower [31,32,33].

### Eligibility criteria for individuals delivering the interventions

The interventions were delivered synchronously by licensed physiotherapists with experience in PD rehabilitation. Before trial delivery, all physiotherapists received standardized training provided by the principal investigator. Training covered the structure and content of both intervention arms, standardized verbal instructions, safety procedures for remote delivery, and procedures to ensure protocol fidelity.

To confirm consistency across therapists, each physiotherapist completed supervised simulations with volunteer patients before delivering the intervention to trial participants. Only physiotherapists who demonstrated adequate adherence to the intervention protocol and uniformity in delivery were authorized to conduct the sessions.

### Intervention and comparator

All assessments and interventions were conducted remotely through real-time video calls. Both groups received 10 synchronous training sessions over six weeks, with two sessions per week during the first four weeks and one session per week during the final two weeks. Each session lasted 45-60 minutes (Table 1). The full intervention protocol, including session-by-session content, standardized mental practice scripts, physical practice exercises, and visual materials, has been described in the published preprint protocol (https://www.medrxiv.org/content/10.1101/2025.04.28.25326452v1.article-metrics) and is summarized in the Supplementary Material.

**Table 1.** Summary of the experimental and control interventions.

| Component | Experimental group | Control group |
| --- | --- | --- |
| Delivery mode | Remote, synchronous video calls | Remote, synchronous video calls |

| **Component** | **Experimental group** | **Control group** |
| --- | --- | --- |
| Total dose | 10 sessions over 6 weeks | 10 sessions over 6 weeks |
| Session duration | 45-60 min | 45-60 min |
| Shared education | FOG education in session 1 | FOG education in session 1 |
| Specific non-PP component | MP based on DNI | Seated upper-limb stretching |
| PP | Same PP protocol as CG | Same PP protocol as EG |
| PP content | Ankle mobility, weight shifting, speed changes, obstacle negotiation, dual-task walking | Same as EG |
| Progression | Increasing gait and cognitive-motor complexity | Same PP progression |
| Main contrast | MP/DNI + PP | Stretching + PP |

Legend: The table presents the main components of the two intervention arms, including delivery mode, total dose, session duration, shared educational content, specific non-physical practice components, physical practice content, progression, and the main contrast between groups. Both groups received the same remotely delivered physical practice protocol and differed only in the additional non-physical practice component: mental practice based on Dynamic Neuro-Cognitive Imagery principles in the experimental group and seated upper-limb stretching in the control group. CG = control group; DNI = Dynamic Neuro-Cognitive Imagery; EG = experimental group; FOG = freezing of gait; MP = mental practice; PP = physical practice.

## Procedures common to both groups

Before the first assessment and intervention session, all participants received standardized instructions on how to prepare the home environment for safe remote participation. They were instructed to ensure a stable internet connection, position the device to allow adequate visual and auditory communication with the physiotherapist, use a stable chair for seated tasks, and clear an unobstructed area of approximately two meters for standing and walking exercises. Participants were also instructed to remove potential hazards, wear non-slip footwear, and keep a stable support surface nearby. When necessary, the presence of a caregiver or family member was requested to support safety during standing and walking tasks.

In the first session, all participants received a brief structured educational module on FOG. This module addressed the definition of FOG, common internal and external triggers, functional consequences, and compensatory strategies to recognize, anticipate, and manage FOG episodes. After this shared educational component, participants were familiarized with the specific procedures of their allocated intervention.

The physical practice component was identical in both groups and consisted of functional exercises targeting gait-related abilities. Exercises included ankle dorsiflexion, anteroposterior and lateral weight shifting, short-distance walking with speed changes, obstacle negotiation, and walking under cognitive or motor dual-task conditions. Tasks were progressively organized to increase complexity over the intervention period. Before each task, the physiotherapist demonstrated the exercise, verified participant understanding, and checked environmental safety. During task execution, the physiotherapist monitored performance by video and provided real-time verbal feedback and corrections when needed.

## Experimental intervention: mental practice plus physical practice

The experimental intervention combined mental practice, supported in Dynamic Neuro-Cognitive Imagery principles, with physical practice. In the remaining nine sessions, each session included two 10-minute mental practice blocks and two 10-minute physical practice blocks, totaling 20 minutes of mental practice and 20 minutes of physical practice.

Mental practice was performed in a seated position, with participants instructed to use a first-person perspective and to minimize overt movement. Standardized verbal scripts were used to guide vivid, kinesthetic, and context-specific imagery of gait-related situations commonly associated with FOG. The imagery scenarios progressively targeted ankle mobility, postural adjustments, lateral weight shifting, walking under time pressure, obstacle negotiation, and walking under dual-task conditions. Visual materials corresponding to each imagery scenario were presented before the respective mental practice blocks to support task comprehension and imagery vividness.

## Control intervention: stretching plus physical practice

The control intervention was designed to match the experimental intervention in duration, therapist contact, remote delivery, and exposure to physical practice. Participants in the control group performed the same physical practice blocks as the experimental group. However, the mental practice blocks were replaced by seated stretching exercises for the scapular region, shoulders, elbows, wrists, and fingers. These exercises were selected as low-intensity activities not specifically designed to target gait performance or FOG.

Thus, the only planned difference between groups was the inclusion of mental practice grounded in Dynamic Neuro-Cognitive Imagery principles in the experimental intervention.

## Intervention fidelity

Intervention fidelity was supported by standardized protocols, structured verbal instructions, predefined progression of task complexity, and prior training of the physiotherapists delivering the sessions. Details of the intervention content, progression, standardized scripts, and visual materials are provided in Supplementary Tables 3,4,5,6 and Supplementary Figure 7.

**Table 2.** Baseline demographic and clinical characteristics of the participants.

|  | CG (n=23) | EG (n=20) | p-value |
| --- | --- | --- | --- |
| **Age (years)** | 55.7 ± 10.3 | 55.7 ± 8.2 | 0.993 |
| **Education (years)** | 16.0 ± 3.3 | 14.1 ± 4.9 | 0.159 |
| **H&Y** | 3.29 ± 0.56 | 3.05 ± 0.62 | 0.220 |
| **LEDD (mg)** | 840.5 ± 339.0 | 776.3 ± 395.2 | 0.584 |

Legend: Data are expressed as mean and standard deviation. H&Y (Hoehn & Yahr); LEDD (Levodopa equivalent daily dose).

**Table 3.** Description of the Experimental Intervention.

| Key points of Gait | Session | Experimental Intervention |
| --- | --- | --- |
| Ankle Mobility | 1/2/3/4/6/10 | **Procedure:** Participants will be asked to imagine dorsiflexing their ankle to release a foot that is held to the floor by a strong Velcro strap. They will be instructed to focus on the sensation of the foot being held to the floor, similar to that which one might experience during a freezing episode. They will then be instructed to feel the foot releasing from the floor by performing an appropriate dorsiflexion movement from the toes to the heel. Instructions will be reinforced with the aid of Figure 1a.<br>**Instructions:** "Imagine that you are sitting on a sturdy chair, with your feet visible. You are wearing shoes with strong Velcro on the soles, firmly attached to the floor. Feel the sensation of your foot stuck to the floor as if you are experiencing a freezing episode. Now, to release your foot, begin to move the tip of your right foot, feeling the Velcro gradually coming off. Concentrate on the sensation in your ankle as you do this. Feel your foot slowly detach, from your toes to your heel. Once your foot is free, place it back on the floor. Let's start again, this time with your left foot." |
| Postural adjustments | 1/2/3/10 | **Procedure:** Participants will be instructed to imagine themselves standing upright while holding a soft ice cream cone with both hands. They will be asked to vividly visualize and kinesthetically sense the weight of the ice cream cone gradually shifting forward. Upon perceiving this forward shift, participants will be guided to mentally realign their posture by moving their trunk backward to prevent the ice cream from toppling over. The focus will be on maintaining the correct postural alignment by adjusting the trunk backward and evenly redistributing body weight across both feet. These instructions will be reinforced through the use of a visual aid (Figure 1b) illustrating the correct posture and intended movement strategy.<br>**Instructions:** "Imagine that you are standing, holding a soft ice cream cone close to your chest with both hands. Now, picture the ice cream starting to tilt forward as your body gently leans forward, as if you were getting tired. To keep the ice cream from falling, slowly bring your upper body back, lifting your chest and straightening your posture. Feel your body moving slightly forward and then backward, as if you are keeping the ice cream safe. Try to balance your weight evenly between both feet. Let's begin again, this time paying even more attention to the feeling of your body coming back into a balanced and steady posture." |
| Lateral weight shifting | 3/4/5/6 | **Procedure:** participants will be instructed to imagine a common situation at home where FOG typically occurs. They will be asked to visualize themselves in their bedroom, preparing to walk toward the living room. As they approach the doorway, they suddenly feel their feet stuck to the floor, unable to take a step, while their body begins leaning forward. Participants will be guided to stay calm, shift their body weight backward to realign their posture, and then perform rhythmic lateral weight-shifting movements, transferring their weight from the right foot to the left, and vice versa. Once they mentally perceive the sensation of the feet "releasing" from the floor, they will imagine taking a step forward to resume walking. This strategy incorporates frequent FOG triggers encountered at home, such as turning and gait initiation. Visual reinforcement will be provided using Figure 1c.<br>**Instructions:** "Imagine that you are at home, in your bedroom, getting ready to walk toward the living room. As you reach the doorway, you suddenly feel your feet stuck to the floor, you have frozen! Your body starts leaning forward, but you cannot move. Stay calm. Now, slowly shift your weight backward, bringing your body back into balance. Gently move your weight from your right foot to your left foot, and then back again. Feel how, with each shift, your feet start to loosen from the floor. When you feel ready, imagine taking a strong, confident step forward to continue walking. Let's go through it again. This time, focus even more on how gently shifting your weight from one foot to the other helps your body unlock from the freezing and makes it easier to take the next step." |
| Walking under pressure | 5/6/7/8 | **Procedure:** Participants will be instructed to imagine themselves walking as fast as possible to a bus stop to catch a bus that was about to depart. To increase the complexity of the situation, participants were warned that if they did not reach the bus, they would have to wait over an hour for the next one. The focus remained on maintaining attention to the key components of gait while managing the pressure of time and emotional excitement. The instructions were reinforced using figure 1d.<br>**Instructions:** "Imagine that you were walking down the sidewalk toward the nearest bus stop. When you looked back, you saw the bus you needed approaching! The stop is still at the end of the block, and you need to walk faster. Take longer, quicker steps to catch it! Feel the impact of your feet on the firm ground and the movement as they detach from the ground with each step. One step after the other, right, left. Take longer, quicker steps, the bus is coming! Let's go through it again. This time, pay special attention to how taking longer steps helps you stay in control, even under pressure." |
| Walking under unpredictable obstacles | 7/8/9 | **Procedure:** Participants will be asked to imagine themselves walking down a busy street, navigating between expected and unexpected obstacles. The physiotherapist guided them to manage their attention between environmental demands and key gait components. The focus remained on promoting adaptive gait strategies in response to environmental challenges. The instructions were reinforced using figure 1e.<br>**Instructions:** "Imagine that you are walking through a busy market, surrounded by colorful fruit and vegetable stalls. You can smell the fresh food and hear the vendors calling out their prices. As you walk, feel your body moving steadily, transferring weight from one leg to the other, with your arms swinging naturally at your sides. Suddenly, a man almost bumps into you. A woman with a cart passes very close to your feet. Then, someone drops a bag of oranges right in front of you! Stay calm. Carefully step to the side - first to the right, then to the left, to avoid the obstacles. Keep your balance as you move around them. After you pass through, you try to turn toward the tomato stall, but suddenly your feet feel stuck to the floor - you have frozen, and your body starts leaning forward. Remember your strategy. First, shift your weight backward to realign your body and prevent falling forward. Bring your chest up and move your center of gravity back. Then, gently start transferring your weight from your right foot to your left foot, and back again. Feel the shift from side to side, slowly and steadily. As you do this, notice how your feet begin to loosen from the floor. When you feel your feet free again, take a confident step forward and continue walking. Let's go through it again, this time paying close attention to both parts of the strategy: first bringing your weight back to regain balance and then shifting side to side to release your feet and move forward." |
| Walking under dual-task | 7/8/9/10 | **Procedure:** Participants will be instructed to imagine themselves inside a pharmacy, moving through the aisles to search for specific items they had memorized before the start of the training. This scenario integrated goal-directed movement, navigation, and working memory, requiring attention to both spatial orientation and task execution. The focus remained on maintaining gait performance while managing cognitive or motor demands simultaneously. The instructions were reinforced using figure 1f.<br>**Instructions:** "Imagine that you are walking to the pharmacy on a rainy day, holding an umbrella in one hand. As you walk, you mentally review your shopping list: dipyron, losartan, omeprazole, eye drops, cotton, and gauze. Feel the ground under your feet with each step. Right foot left foot - transferring your weight steadily from one leg to the other. When you arrive at the pharmacy, you gently close your umbrella and start walking between the shelves. Stay calm and focused. Move carefully and steadily while searching for the items on your list. Notice how your body keeps its balance even as your mind works to remember and find what you need. Now, mentally recall your list again: what were the medications and items you needed to find? Let's go through it one more time, paying attention to how you keep walking safely while thinking about your tasks." |

Legend: The table presents each of the key gait components emphasized, the sessions in which they will be addressed, the procedure will be adopted for their execution, and the structured and standardized verbal instructions provided to participants during Mental Practice (mental practice). FOG = Freezing of Gait.

**Table 4.** Description of physical Practice (PP).

| Key points of Gait | Session | Experimental Intervention |
| --- | --- | --- |
| Ankle mobility | 1/2/3/4/6/10 | **Procedure:** Participants will be instructed to perform ankle mobility exercises aimed at improving dorsiflexion range of motion, both in seated and standing positions.<br>**PP:** Ankle dorsiflexion exercises will include: (1) repetitive ankle dorsiflexion movements performed both unilaterally and bilaterally simultaneously; (2) seated foot mobilization by rolling a water bottle under the plantar surface; and (3) closed kinetic chain dorsiflexion exercises performed in a standing position with an extended anteroposterior base. For the closed kinetic chain task, participants will be instructed to flex the knee of the front lower limb while maintaining heel contact with the ground to facilitate active ankle dorsiflexion. |
| Postural adjustments | 3/4/5/6 | **Procedure:** Participants will be instructed to perform repetitive anteroposterior weight-shifting movements in the standing position. The objective of this exercise is to promote dynamic postural control by encouraging controlled forward and backward body displacements while standing.<br>**PP:** Participants will perform repetitive anteroposterior body movements while standing, varying the base of support between a neutral stance and an extended anteroposterior stance. Variations in movement, amplitude and speed will also be incorporated to progressively challenge balance and mobility. Movements will be performed under supervision to ensure correct execution and safety. |
| Lateral weight shifting | 3/4/5/6 | **Procedure:** Participants will be instructed to perform lateral weight-shifting exercises while standing, with the objective of promoting mediolateral postural control and balance adjustment.<br>**PP:** Exercises will involve repetitive latero-lateral body movements performed in a standing position, with variations in knee positioning, including execution with semi-flexed knees and with extended lower limbs. Alternating single-leg support tasks will also be incorporated, along with progressive variations in movement amplitude and speed to increase the complexity of the task. |
| Walking under pressure | 5/6/7/8 | **Procedure:** Participants will be instructed to walk a short distance while responding to verbal commands indicating sudden changes in gait speed, with the objective of promoting adaptability and dynamic gait control.<br>**PP:** Exercises will involve walking with sudden changes in speed, transitioning from faster to slower paces, incorporating sudden stops, and executing wider steps as directed. |
| Walking under unpredictable obstacles | 7/8/9 | **Procedure:** Participants will be instructed to walk a short distance while negotiating environmental obstacles, aiming to improve attentional control and adaptability during gait.<br>**PP:** Exercises will involve walking while negotiating obstacles (e.g., toilet paper rolls) arranged in different configurations, including zigzag paths, narrow passages, and alternating lateral deviations (i.e., passing around obstacles from the left or right |

side).
| Key points of Gait | Session | Experimental Intervention |
| --- | --- | --- |
| Walking under dual-task | 7/8/9/10 | **Procedure:** Participants will be instructed to walk a short distance while performing simultaneous cognitive or motor secondary tasks, with the objective of enhancing gait performance under dual-task conditions.<br>**PP:** Exercises will involve walking while performing dual tasks, such as solving mental calculations, spelling words backward, listing words starting with a specific letter, reciting the months of the year in reverse order, recalling a shopping list, or carrying a half-filled glass of water. |

Legend: The table presents each of the key gait components and the sessions in which they will be addressed, the procedure will be adopted for their execution, and the structured and standardized instructions provided to participants during PP (physical practice). FOG = Freezing of Gait.

**Table 5.**
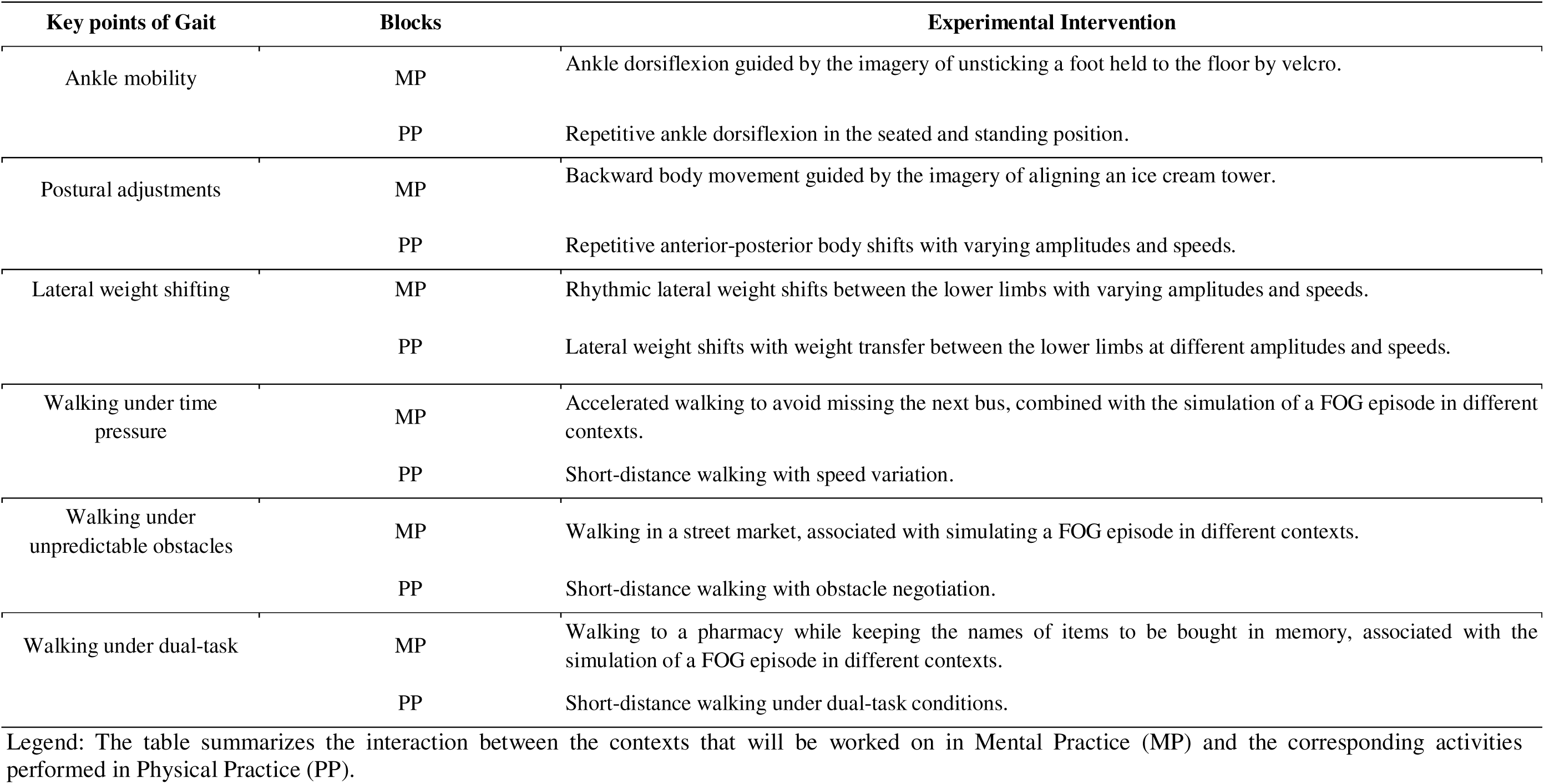
Description of the interaction between Mental Practice (MP) and Physical Practice (PP)

| Key points of Gait | Blocks | Experimental Intervention |
| --- | --- | --- |
| Ankle mobility | MP | Ankle dorsiflexion guided by the imagery of unsticking a foot held to the floor by velcro. |
|  | PP | Repetitive ankle dorsiflexion in the seated and standing position. |
| Postural adjustments | MP | Backward body movement guided by the imagery of aligning an ice cream tower. |
|  | PP | Repetitive anterior-posterior body shifts with varying amplitudes and speeds. |
| Lateral weight shifting | MP | Rhythmic lateral weight shifts between the lower limbs with varying amplitudes and speeds. |
|  | PP | Lateral weight shifts with weight transfer between the lower limbs at different amplitudes and speeds. |
| Walking under time pressure | MP | Accelerated walking to avoid missing the next bus, combined with the simulation of a FOG episode in different contexts. |
|  | PP | Short-distance walking with speed variation. |
| Walking under unpredictable obstacles | MP | Walking in a street market, associated with simulating a FOG episode in different contexts. |
|  | PP | Short-distance walking with obstacle negotiation. |
| Walking under dual-task | MP | Walking to a pharmacy while keeping the names of items to be bought in memory, associated with the simulation of a FOG episode in different contexts. |
|  | PP | Short-distance walking under dual-task conditions. |

Legend: The table summarizes the interaction between the contexts that will be worked on in Mental Practice (MP) and the corresponding activities performed in Physical Practice (PP).

**Table 6.**
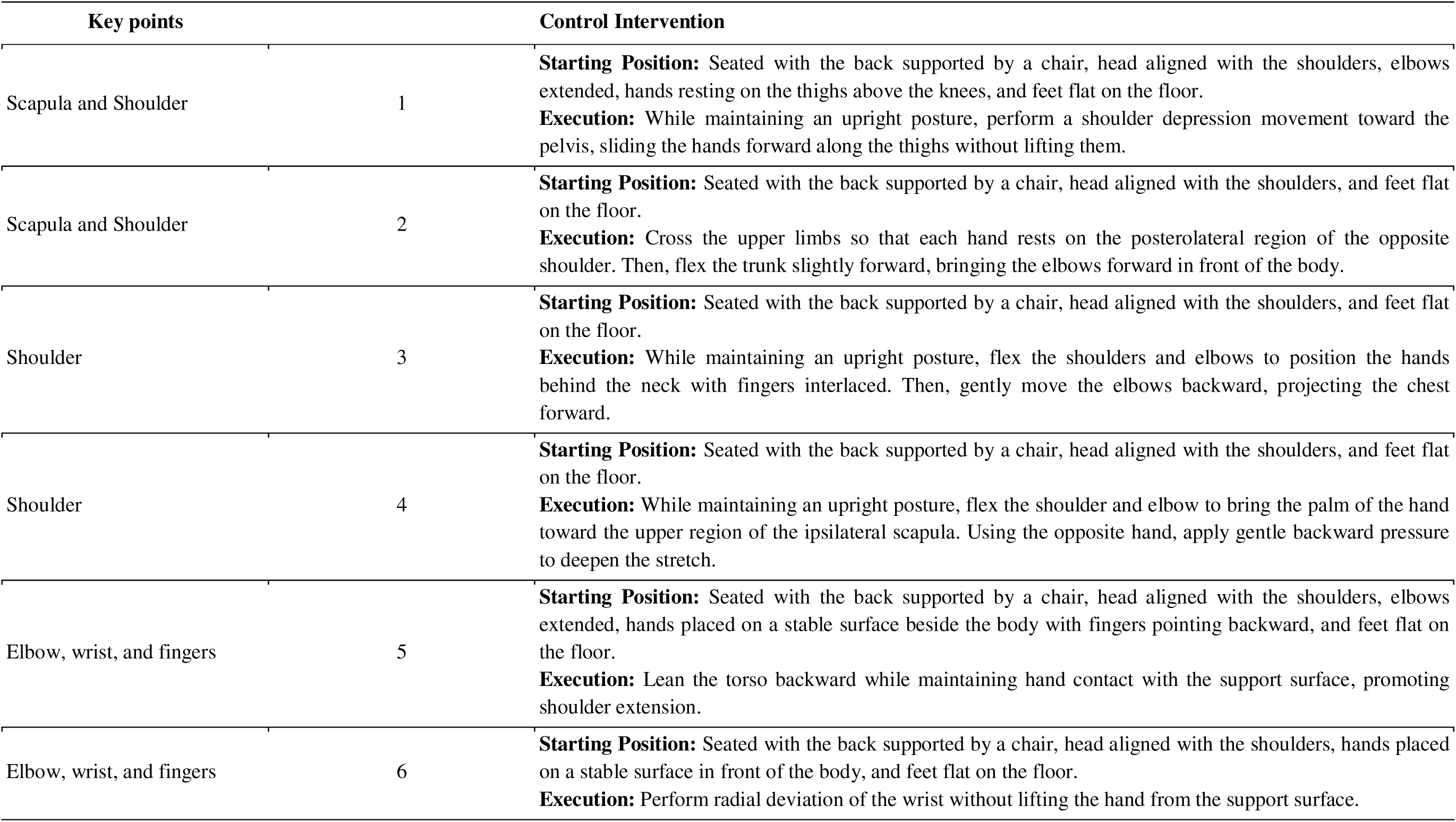
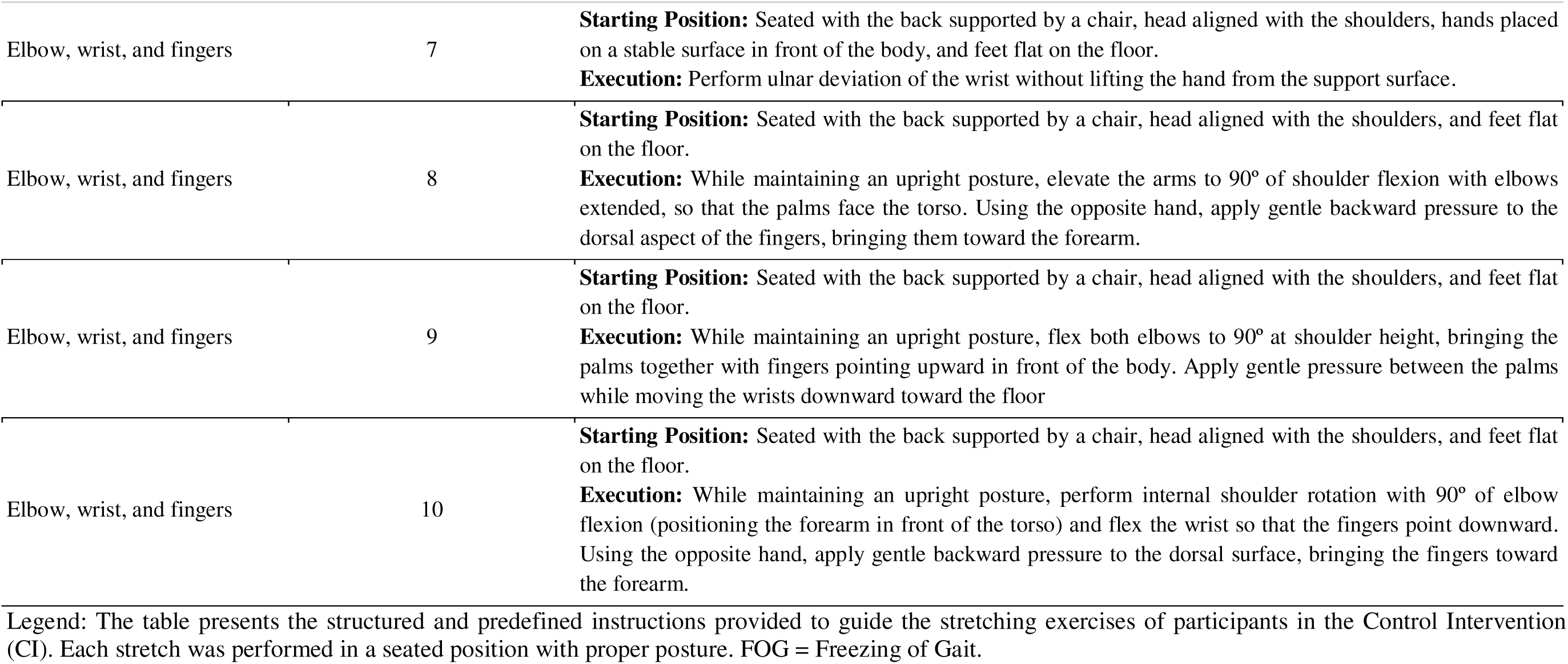
Description of the Control Intervention (CI).

### Outcomes

Participants in both groups were assessed at three time points: baseline (before intervention), immediately after the intervention, and 30 days after completion of the intervention as a follow-up. Demographic information, including date of birth, age, sex, educational level, and occupation, was collected at baseline.

## Primary outcome

The primary outcome was performance during the Rapid Turns Test, a standardized provocative task designed to elicit FOG during rapid turning. This outcome was quantified using two prespecified task-derived metrics: total test duration and percentage of time spent with FOG during the test.

The Rapid Turns Test was selected because rapid turning is one of the most consistent triggers of FOG episodes and the test has been reported as a reliable, valid, and sensitive procedure for identifying FOG [34,35]. Participants were instructed to repeatedly turn around their own axis, alternating directions, as quickly and continuously as possible until instructed to stop [36].

Before the test, the evaluator verified that the home environment was suitable for safe execution. Participants, and when available a caregiver or family member, were instructed to remove obstacles and rugs, ensure adequate lighting, and remain close to a stable support surface, such as a wall, countertop, or sturdy chair. The evaluator explained the task, demonstrated the procedure, confirmed participant understanding, and ensured that the participant felt safe before starting the test.

The test was initiated after the verbal cue “Now.” Throughout the task, the evaluator maintained a clear video view of the participant and supervised performance to ensure safety.

The task was video recorded for subsequent analysis.

Total test duration was defined as the time, in seconds, required to complete the Rapid Turns Test. FOG episodes were visually scored from the video recordings. The percentage of time spent with FOG was calculated as:

**Percentage of time spent with FOG = total duration of FOG episodes during the test × 100 / total test duration.**

## Secondary outcomes

### I) Self-reported FOG severity by NFOG-Q

Participants completed the self-reported questionnaire while seated. The NFOG-Q assesses clinical characteristics of FOG and its impact on quality of life. Total scores range from 0 to 28, with higher scores indicating greater FOG severity and impact [28].

### **II)** Movement Disorders Society - Unified Parkinson’s Disease Rating Scale (UPDRS)

The UPDRS is a widely used instrument for assessing and monitoring the progression of PD. It comprises 42 items divided into four parts: Part I (non-motor aspects of daily living), Part II (motor aspects of daily living), Part III (motor examination), and Part IV (motor complications). Participants completed Part II while seated. This section consists of 13 items, each scored from 0 to 4, with higher scores indicating greater impairment in motor aspects of daily living [37].

### **III)** Mobility domain by Parkinson’s Disease Questionnaire–39 (PDQ-39)

Participants completed the questionnaire while seated. The PDQ-39 consists of 39 items distributed across eight domains: mobility (10 items), activities of daily living (6 items), emotional well-being (6 items), stigma (4 items), social support (3 items), cognition (4 items), communication (3 items), and physical discomfort (3 items). Each item is rated on a five-point Likert scale (never, rarely, sometimes, often, always). Domain and total scores range from 0 to 100, with lower scores indicating better perceived quality of life [38]. Participants completed in this study only the mobility domain while seated.

### **IV)** Global cognitive capacity by T-MoCA

The T-MoCA was administered while participants were seated. This instrument evaluates six cognitive domains, including naming, memory, attention, language, abstraction, and orientation, and does not require paper, pencil, or visual stimuli. The maximum possible score is 22 points [29].

#### Harms

Adverse events were defined as any undesirable medical, motor, or non-motor event occurring during the intervention period, regardless of whether it was considered related to the intervention. Events specifically monitored during sessions included falls, near falls, pain, excessive fatigue, dizziness, shortness of breath, motor fluctuations, emotional discomfort, or any other complaint requiring session interruption or modification.

Harms were systematically monitored by the physiotherapist during each remote training session. At the beginning and end of each session, participants were asked about adverse events, clinical changes, or difficulties since the previous session. Any event reported by the participant or observed by the physiotherapist was recorded, including its timing, description, severity, need for session modification or discontinuation, and presumed relationship to the intervention. Serious adverse events were defined as events resulting in injury, urgent medical care, hospitalization, or inability to continue the intervention.

#### Sample size

The Rapid Turn Test and the percentage of time spent with FOG during the test were selected as primary performance-based outcomes because rapid turning is one of the most consistent triggers of FOG and provides an objective assessment of FOG during a standardized provocative gait task. Although the Rapid Turn Test has been used in previous studies to provoke or identify FOG, no previous randomized controlled trial was identified that provided adequate parameters for sample size estimation using Rapid Turn Test completion time or percentage of time spent with FOG as primary outcomes. Therefore, the sample size calculation was based on a clinically related measure of FOG severity.

The sample size was estimated using data from a previous randomized controlled trial in people with PD and FOG that investigated action observation combined with physiotherapy and reported a significant group × time interaction for FOG severity [39]. Based on the reported interaction effect, F(2,59) = 9.49, the estimated partial eta squared was 0.24, corresponding to a Cohen’s f of approximately 0.57. Assuming a two-sided significance level of 0.05, statistical power of 80%, two groups, three repeated assessments, and an estimated correlation of 0.50 between repeated measures, the required sample size was estimated at 12 participants per group. To account for potential losses to follow-up, the target sample size was increased to at least 15 participants per group.

### Interim analyses and stopping guidelines

No interim analysis was planned or conducted. The trial did not include predefined stopping rules, as the intervention was low risk, delivered remotely under physiotherapist supervision, and had a short duration.

## Randomization

### Sequence generation

The random allocation sequence was generated by an independent researcher who was not involved in participant recruitment, intervention delivery, outcome assessment, or data analysis. Participants were stratified according to cognitive performance assessed by the T-MoCA and then randomized within each stratum to the experimental group or control group in a 1:1 allocation ratio. The random sequence was generated using Microsoft Excel.

### Type of randomization

A stratified randomization procedure was used to promote balance between groups in cognitive performance, given the relevance of cognition to motor imagery, gait control, and FOG. No blocking procedure was used.

### Allocation concealment mechanism

Allocation concealment was maintained by restricting access to the randomization sequence. The allocation list was kept by the independent researcher who generated the sequence and was not accessible to the outcome assessor. Group assignment was revealed only after completion of baseline assessment and participant enrolment.

### Implementation

Participants were enrolled and completed baseline assessments before randomization. After baseline assessment, the independent researcher communicated group assignment to the physiotherapist responsible for intervention delivery. Participants were then allocated to either the experimental group, which received mental practice grounded in Dynamic Neuro-Cognitive Imagery plus physical practice, or the control group, which received seated stretching plus the same physical practice protocol.

The outcome assessor did not participate in intervention delivery and remained blinded to group allocation throughout data collection. Participants were instructed not to disclose any information about their intervention content during reassessments.

## Blinding

### Who was blinded

The outcome assessor responsible for baseline, post-intervention, and follow-up assessments was blinded to group allocation. Participants and physiotherapists delivering the intervention could not be blinded because of the nature of the intervention. The assessor became aware of group allocation only after completion of all assessments and initiation of the final data analysis.

### How blinding was achieved

Blinding was achieved by separating the roles of intervention delivery and outcome assessment. The physiotherapists responsible for delivering the interventions did not participate in any assessment procedures, and the outcome assessor did not have access to the randomization list or intervention records during data collection.

Both interventions were delivered remotely, followed the same schedule, frequency, session duration, therapist contact, and progression of physical practice. Both groups received the same physical practice protocol and differed only in the non-physical practice component: mental practice grounded in Dynamic Neuro-Cognitive Imagery in the experimental group and seated upper-limb stretching in the control group. This structural similarity was intended to reduce the risk of unblinding of the assessor during participant interactions.

### Statistical methods Statistical methods

Baseline demographic and clinical characteristics were compared between groups to describe sample comparability before the intervention. The distribution of continuous variables was assessed using the Shapiro–Wilk test and visual inspection of the data. Variables with approximately normal distribution were compared using independent-samples Student’s t tests, whereas non-normally distributed or ordinal variables were compared using Mann-Whitney U tests. Categorical variables, when applicable, were compared using chi-square or Fisher’s exact tests.

Intervention effects on the primary task-derived metrics from the Rapid Turns Test and on secondary outcomes were analyzed using permutation-based repeated-measures analysis of variance with 10,000 permutations. The model included group, time, and group × time interaction effects, with group as the between-participant factor and time as the within-participant factor. This approach was selected because permutation-based inference is less dependent on parametric distributional assumptions and is more robust for small samples and outcomes with non-normal distributions.

When significant main or interaction effects were identified, they were further explored using Tukey-adjusted post hoc pairwise comparisons. For the primary Rapid Turns Test metrics, interpretation focused on the consistency of findings across total test duration and percentage of time spent with FOG, as both metrics were derived from the same prespecified provocative task and captured complementary aspects of performance.

A linear trend analysis was additionally performed to explore patterns of change in the percentage of time spent with FOG across the three assessment points within each group. Linear regression coefficients, coefficients of determination (R²), and p values were calculated separately for each group. This analysis was considered complementary and exploratory.

All analyses were performed using Python software, version 3.11. Statistical significance was set at a two-sided alpha level of 0.05.

### Analysis population

All randomized participants were included in the primary and secondary outcome analyses according to their originally assigned groups, following the intention-to-treat principle. Participants with missing post-intervention or follow-up data were retained in the analysis after imputation of missing values.

### Missing data

Missing outcome data occurred because of participant loss after randomization. For the intention-to-treat analysis, missing post-intervention and follow-up values were imputed using group-specific mean imputation for the corresponding outcome and assessment time point. This approach was chosen to retain all randomized participants in the analysis while preserving the original group allocation.

To assess the robustness of the findings to the handling of missing data, an exploratory sensitivity analysis compared the results obtained using group-mean imputation with alternative imputation strategies, including regression imputation and last observation carried forward. The direction and overall pattern of the main findings were examined across these approaches.

### Additional analyses

A prespecified linear trend analysis was conducted to evaluate within-group changes in the percentage of time spent with FOG across baseline, post-intervention, and follow-up assessments. No subgroup analyses were planned or performed. The comparison of missing-data imputation strategies was conducted as an exploratory sensitivity analysis.

## Results

### Participants flow

A total of 64 individuals were screened for eligibility and completed the baseline assessment. Of these, 21 were excluded before randomization: seven did not report FOG, as indicated by a negative response to item 1 of the NFOG-Q; one required an assistive device for ambulation at home; two had insufficient motor imagery ability according to the KVIQ-20; and eleven scored below 12 points on the T-MoCA.

The remaining 43 participants were randomized to the experimental group (n = 20) or the control group (n = 23). During the trial, five participants were lost to follow-up because of absenteeism: two in the experimental group and three in the control group. In the experimental group, one participant was lost after baseline and one after the post-intervention assessment. In the control group, three participants were lost after baseline. Therefore, 38 participants completed the intervention protocol and all planned reassessments: 18 in the experimental group and 20 in the control group.

All 43 randomized participants were included in the intention-to-treat analyses according to their originally assigned groups, with missing post-intervention or follow-up data handled as described in the statistical analysis section.

### Recruitment

Participants were recruited, assessed, and enrolled between May and December 2025. Intervention delivery and follow-up assessments were completed within the same period. The final follow-up assessment was conducted 30 days after completion of the intervention.

### Trial completion

The trial ended as planned after completion of recruitment, intervention delivery, and follow-up assessments. No early stopping criteria were applied, and the trial was not discontinued for safety, feasibility, or efficacy reasons.

## Intervention and comparator delivery

### Concomitant care received during the trial for each group

Participants in both groups continued to receive their usual medical care for PD throughout the trial. In accordance with the eligibility criteria, no participant engaged in concurrent physiotherapy specifically targeting gait or FOG during the intervention period.

Antiparkinsonian medication regimens remained unchanged from enrolment to follow-up.

### Baseline data

Baseline demographic and clinical characteristics of participants allocated to the experimental group and control group are presented in Table 2. No statistically significant between-group differences were observed at baseline.

Overall, participants were recruited from all five geographic regions of Brazil. Socioeconomic status, classified according to the Brazilian Economic Classification Criteria, ranged from class A to class D–E.

### Numbers analyzed, outcomes and estimation

All 43 randomized participants were included in the primary and secondary outcome analyses according to their originally assigned groups, following data imputation. Of these, 38 participants completed the intervention protocol and all planned assessments: 18 in the experimental group and 20 in the control group.

## Primary outcome: Rapid Turns Test performance

Rapid Turns Test performance was quantified using two prespecified task-derived metrics: total test duration and percentage of time spent with FOG during the test.

## Total test duration

Permutation-based repeated-measures ANOVA for total test duration revealed significant main effects of group and time, as well as a significant group × time interaction (F(2,96) = 4.088, p = 0.0019, ES = 0.81), as shown in Figure 1.

**Figure 1.**
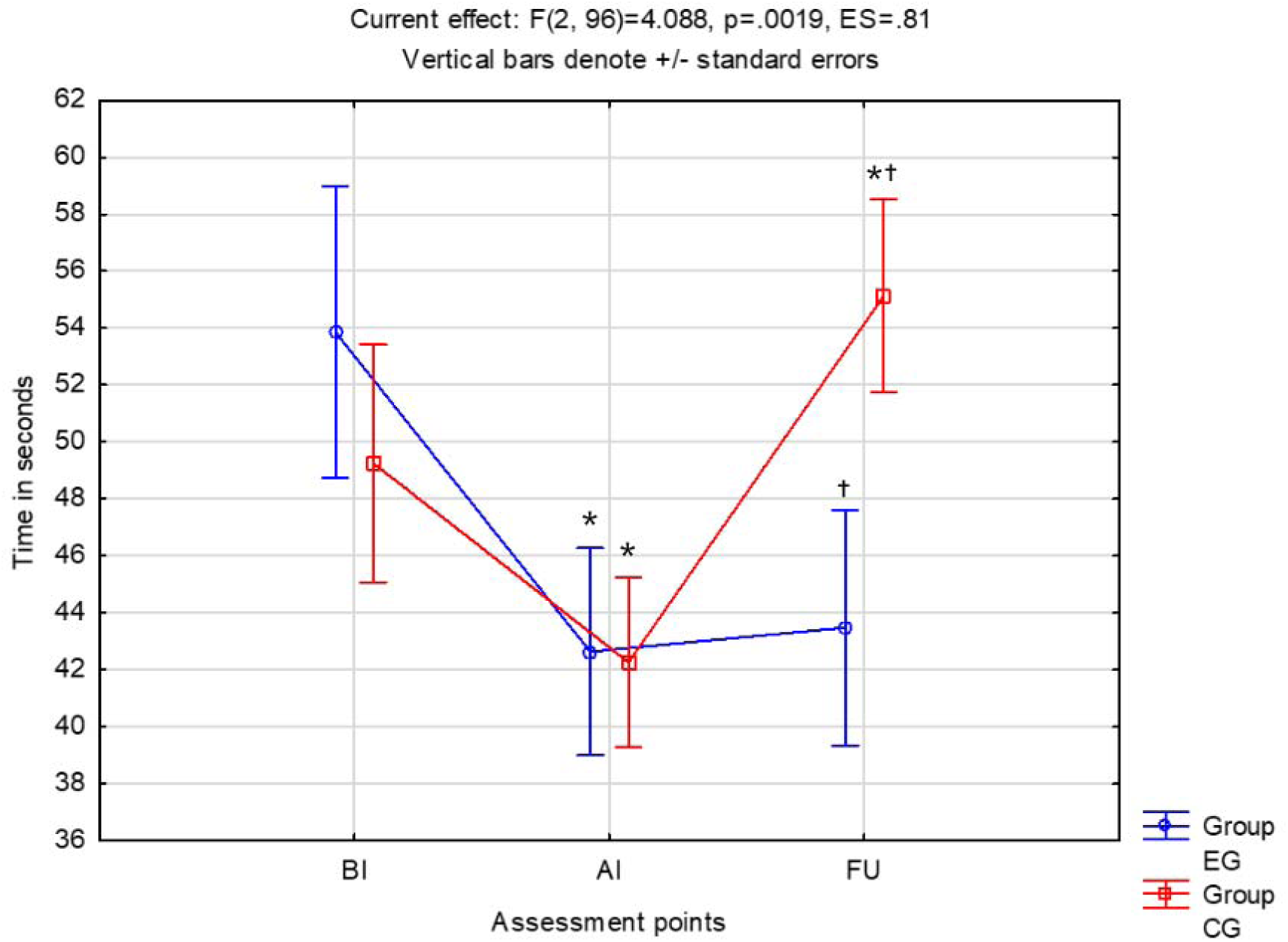
Total test duration during the Rapid Turns Test

Mean ± SD values for the experimental group were 53.9 ± 21.4 seconds before intervention, 42.6 ± 16.4 seconds after intervention, and 43.5 ± 19.2 seconds at follow-up. For the control group, mean ± SD values were 49.7 ± 23.5 seconds before intervention, 44.6 ± 20.5 seconds after intervention, and 55.1 ± 18.1 seconds at follow-up.

Post hoc analyses showed that the experimental group significantly reduced total test duration from before to after intervention, with this improvement maintained at follow-up. In contrast, the control group showed an initial reduction from before to after intervention, followed by a significant increase at follow-up, indicating loss of the initial gain. Between-group comparisons showed no significant differences at before or after intervention, but a significant difference at follow-up, with longer total test duration in the control group than in the experimental group.

## Percentage of time spent with FOG

Permutation-based repeated-measures ANOVA for the percentage of time spent with FOG during the Rapid Turns Test revealed significant main effects of group and time, as well as a significant group × time interaction (F(2,94) = 2.277, p = 0.0108, ES = 0.74), as shown in Figure 2.

**Figure 2.**
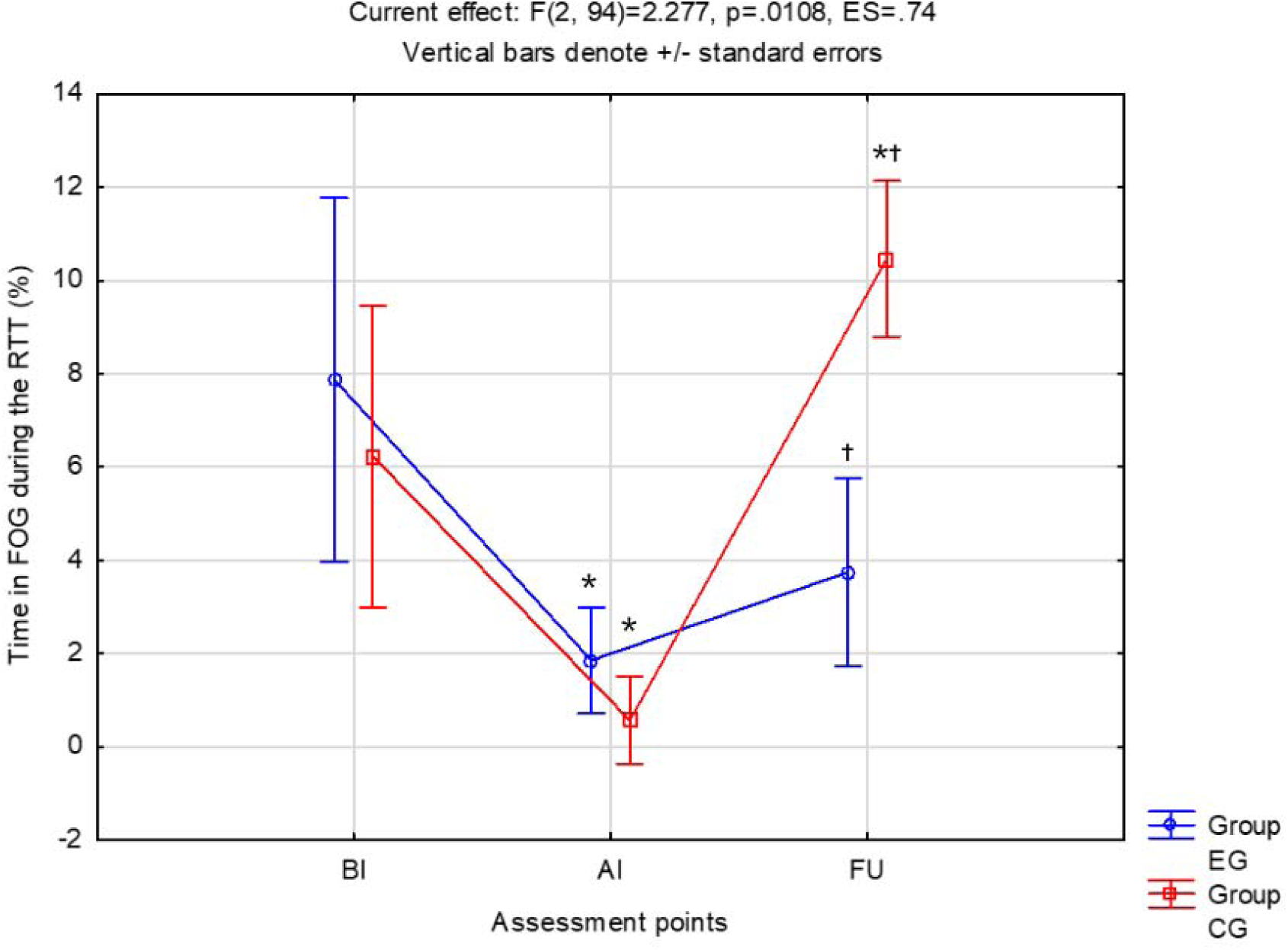
Percentage of time spent with FOG during the Rapid Turns Test.

Mean ± SD values for the experimental group were 7.8 ± 4.2% before intervention, 1.8 ± 1.4% after intervention, and 3.7 ± 1.7% at follow-up. For the control group, mean ± SD values were 6.2 ± 3.8% before intervention, 0.6 ± 0.3% after intervention, and 10.4 ± 1.8% at follow-up.

Post hoc analyses showed that the experimental group significantly reduced the percentage of time spent with FOG from before to after intervention, with no significant change between after intervention and follow-up. In contrast, the control group showed a significant increase at follow-up compared with both before and after intervention. Between-group comparisons showed no significant differences at before or after intervention, but a significant difference at follow-up, with a higher percentage of time spent with FOG in the control group than in the experimental group.

## Secondary outcomes

### Self-reported FOG severity assessed by the NFOG-Q

For self-reported FOG severity assessed by the NFOG-Q, permutation-based repeated-measures ANOVA revealed a significant main effect of time (F(2,102) = 83.100, p < 0.001, ES = 0.99), with no significant main effect of group and no significant group × time interaction, as shown in Figure 3.

**Figure 3.**
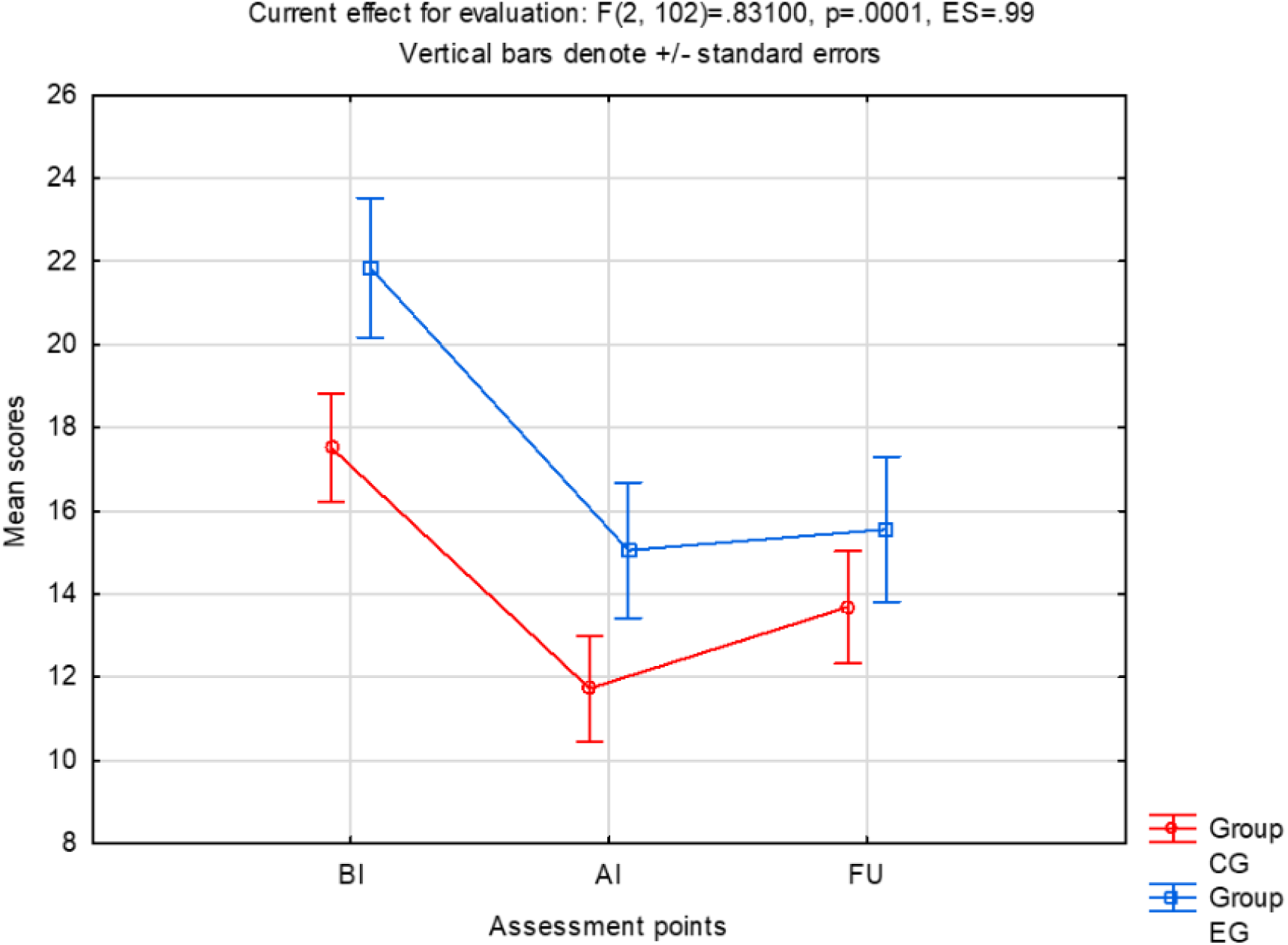
Self-reported FOG severity assessed by the NFOG-Q

Mean ± SD scores for the experimental group were 21.8 ± 6.0 before intervention, 15.0 ± 8.3 after intervention, and 15.5 ± 9.1 at follow-up. For the control group, mean ± SD scores were 17.5 ± 8.2 before intervention, 11.7 ± 6.5 after intervention, and 13.6 ± 6.8 at follow-up.

Tukey-adjusted post hoc comparisons showed significant reductions in NFOG-Q scores from before to after intervention in both groups. No significant differences were observed between after intervention and follow-up within either group, suggesting maintenance of the post-intervention scores. Between-group comparisons did not show significant differences at any assessment point.

## Motor disability by UPDRS

For motor aspects of daily living assessed by MDS-UPDRS Part II, permutation-based repeated-measures ANOVA revealed a significant main effect of time (F(2,102) = 2.610, p = 0.001, ES = 0.98), with no significant main effect of group and no significant group × time interaction, as shown in Figure 4.

**Figure 4.**
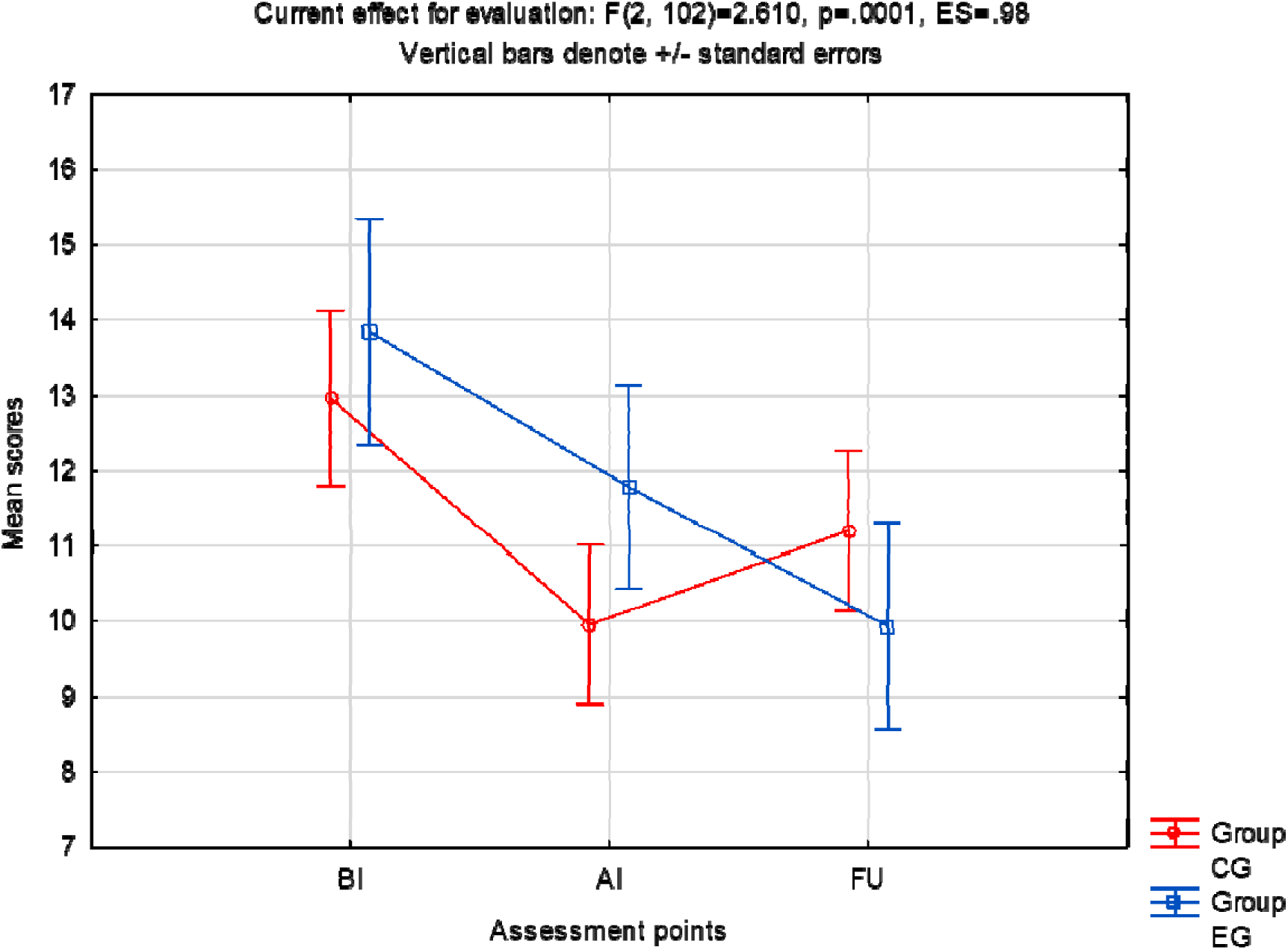
Motor aspects of daily living assessed by MDS-UPDRS Part II.

Mean ± SD scores for the experimental group were 13.8 ± 7.1 before intervention, 11.7 ± 7.1 after intervention, and 9.9 ± 7.4 at follow-up. For the control group, mean ± SD scores were 12.9 ± 6.4 before intervention, 9.9 ± 5.2 after intervention, and 11.2 ± 5.1 at follow-up.

Tukey-adjusted post hoc comparisons did not identify significant pairwise differences between assessment points within either group. No significant differences between-group were observed at any assessment point.

## Mobility domain by PDQ-39

For the PDQ-39 mobility domain, permutation-based repeated-measures ANOVA revealed a significant group × time interaction (F(2,102) = 3.932, p = 0.001, ES = 0.97), indicating different patterns of change between groups across assessments, as shown in Figure 5.

**Figure 5.**
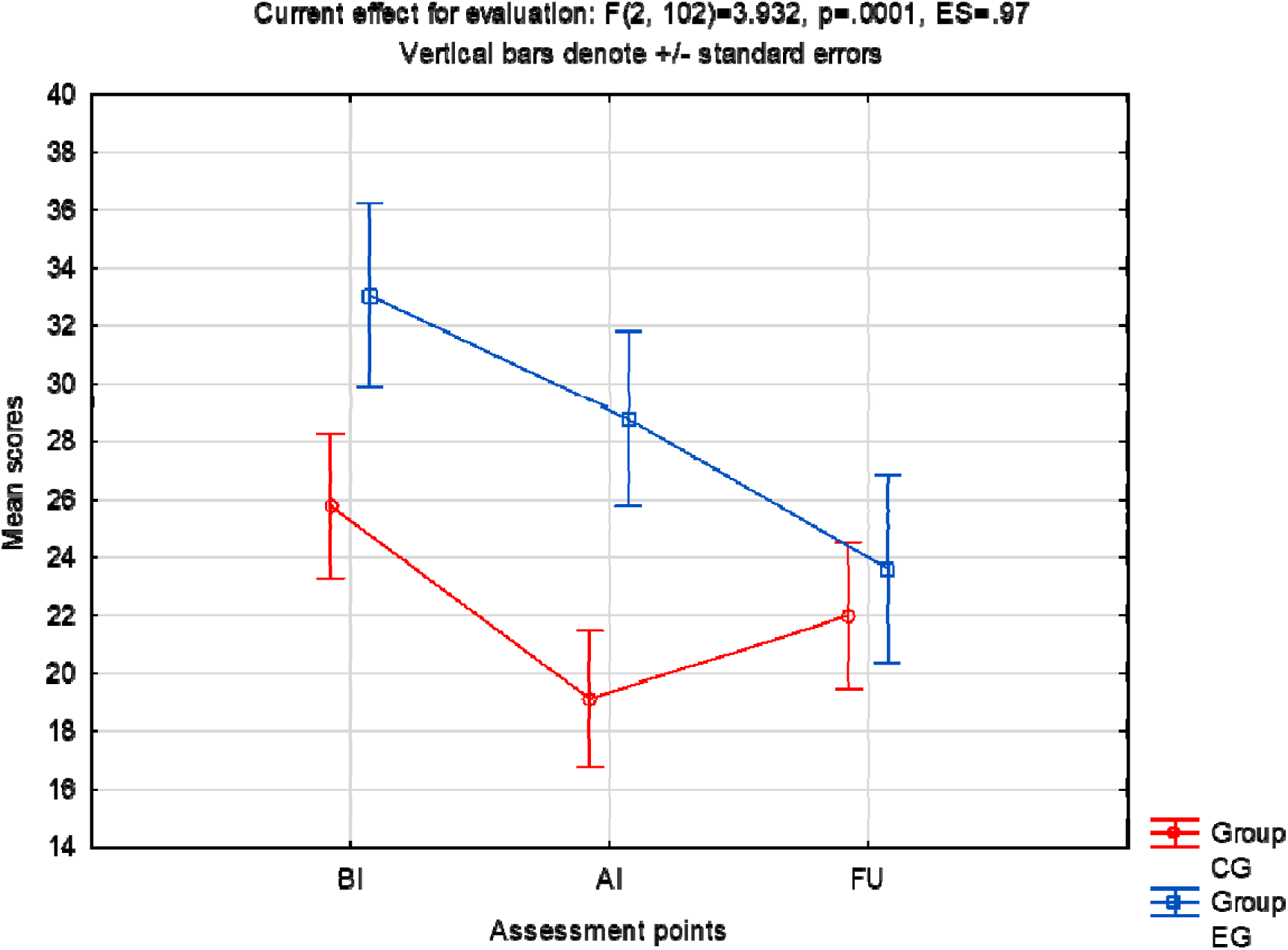
Mobility domain assessed by PDQ-39.

Mean ± SD scores for the experimental group were 33.0 ± 16.1 before intervention, 28.7 ± 14.9 after intervention, and 23.6 ± 17.3 at follow-up. For the control group, mean ± SD scores were 25.7 ± 13.0 before intervention, 19.1 ± 12.6 after intervention, and 22.0 ± 12.5 at follow-up.

Tukey-adjusted post hoc comparisons showed a significant reduction in PDQ-39 mobility scores from before intervention to follow-up in the experimental group (p = 0.0028), consistent with improved mobility-related quality of life. No significant changes were observed in the control group across assessment points. Between-group comparisons were not significant at any assessment point.

## Global cognitive capacity by T-MoCA

For global cognitive performance assessed by the T-MoCA, permutation-based repeated-measures ANOVA revealed significant main effects of group and time, as well as a significant group × time interaction (F(2,102) = 6.70, p = 0.0018, ES = 0.85), as shown in Figure 6.

**Figure 6.**
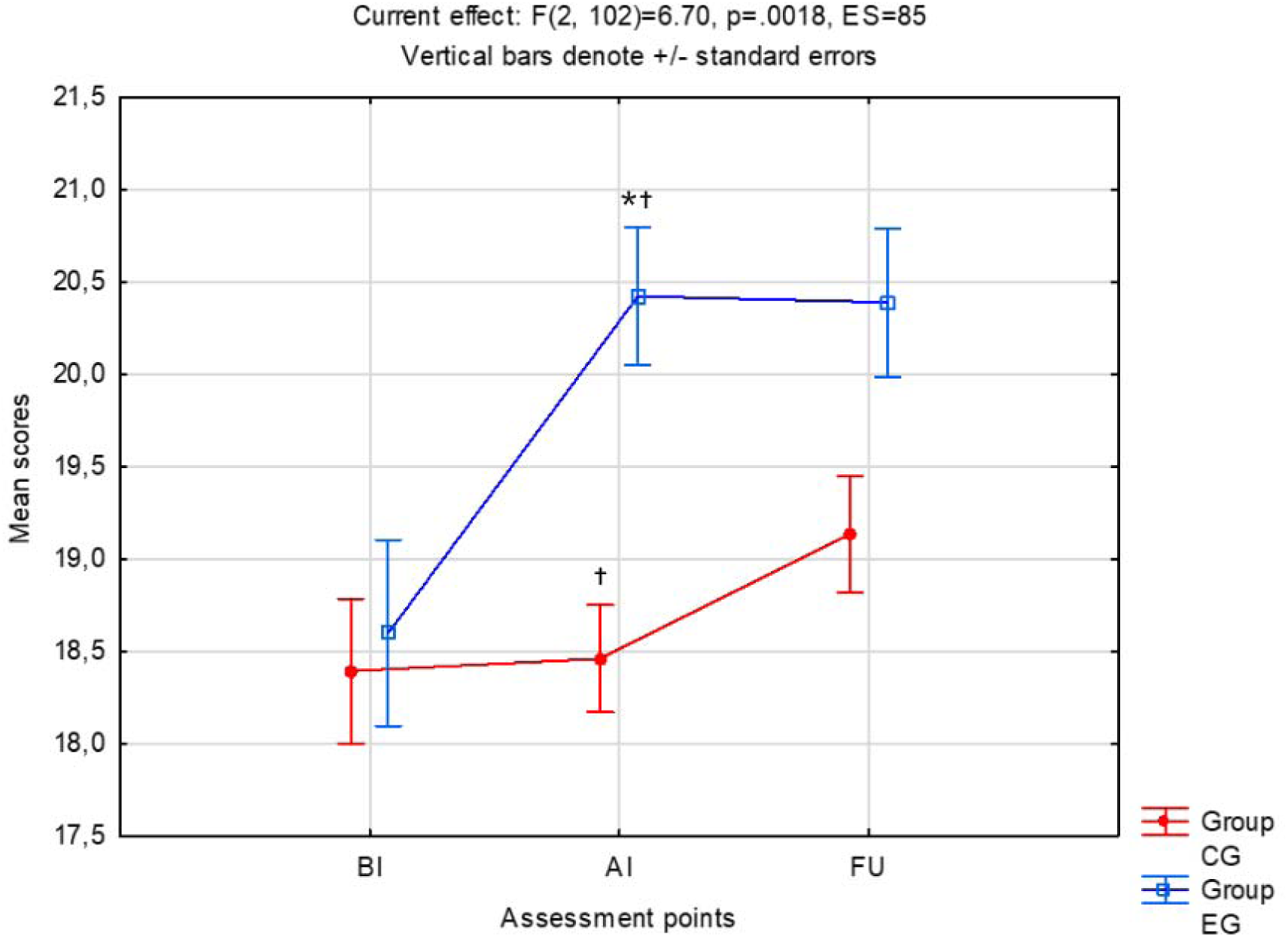
Global cognitive capacity assessed by the T-MoCA.

**Figure 7.**
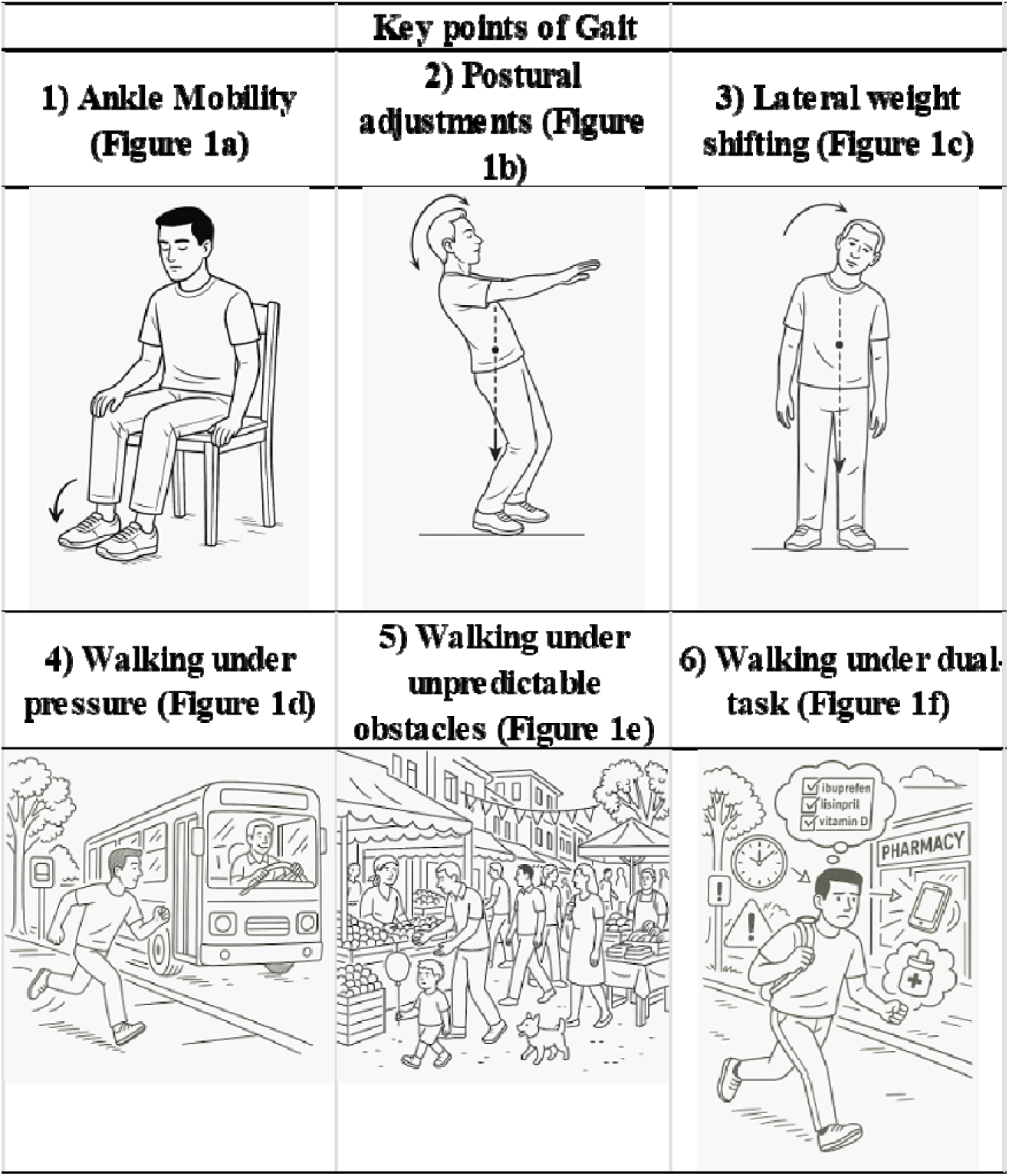
Figures that will be presented to participants to facilitate Mental Practice (MP).

Mean ± SD scores for the experimental group were 18.6 ± 2.7 before intervention, 20.4 ± 1.3 after intervention, and 20.3 ± 1.9 at follow-up. For the control group, mean ± SD scores were 18.3 ± 1.9 before intervention, 18.4 ± 1.8 after intervention, and 19.1 ± 1.7 at follow-up.

Tukey-adjusted post hoc comparisons showed a significant increase in T-MoCA scores from before to after intervention in the experimental group, with no significant change between after intervention and follow-up, suggesting maintenance of the post-intervention score. In the control group, no significant within-group changes were observed across assessment points. Between-group comparisons showed no significant difference before intervention, but a significant difference after intervention, with higher T-MoCA scores in the experimental group than in the control group.

### Harms

No serious adverse events occurred during the intervention period. Two isolated falls were reported, each in a different participant, both during the transition from sitting to standing. In both cases, the participant returned to the seated position without injury, need for medical care, or discontinuation of the intervention.

Other motor and non-motor complaints reported during training, such as motor fluctuations, pain, reduced motivation, or concerns related to participation, were recorded but were not considered intervention-related, as they were present before the trial or reflected usual PD-related fluctuations. No participant discontinued the intervention because of harms.

### Ancillary analyses

No prespecified subgroup analyses were performed. A complementary exploratory linear trend analysis was conducted to examine changes in the percentage of time spent with FOG across before intervention, after intervention, and follow-up within each group.

Sensitivity analyses were also conducted to examine the robustness of the findings to different missing data handling strategies. The results obtained using group-mean imputation were compared with those obtained using regression imputation and last observation carried forward. The overall direction and pattern of the main findings were examined across these approaches.

### Discussion Interpretation

This study addressed an important gap in the rehabilitation of people with PD and FOG: the limited evidence on remotely delivered interventions that combine physical practice with cognitive-motor strategies specifically designed to reinforce internal motor representations and gait control. We investigated whether adding mental practice, grounded in Dynamic Neuro-Cognitive Imagery principles, to physical practice would produce greater benefits than physical practice alone. Overall, the findings suggest that the combined intervention was associated with more favorable trajectories in Rapid Turns Test performance, global cognitive performance, and mobility-related quality of life, whereas both groups showed changes over time in self-reported FOG severity and motor aspects of daily living. This pattern indicates that mental practice may have contributed mainly to outcomes requiring cognitive-motor integration, task-specific gait regulation, and maintenance of gains, rather than producing broad superiority across all clinical measures.

A central finding was that mental practice did not produce clear superiority immediately after the intervention, but was associated with better maintenance of gains after training cessation. This pattern partially supports our initial hypothesis. The short-term improvements observed in both groups suggest that structured physical practice, FOG education, therapist supervision, and repeated exposure to gait-related tasks were sufficient to produce immediate benefits. However, the less stable trajectory observed in the control group after supervised training ended suggests that physical practice alone may have been insufficient to sustain these gains. In contrast, the experimental group showed a more stable pattern over time, particularly in outcomes requiring task-specific gait control. One possible explanation is that mental practice helped participants internalize strategies for managing FOG, allowing them to anticipate FOG episodes, redirect attention to key gait components, reorganize posture, and initiate compensatory strategies without relying exclusively on external feedback from the therapist.

A complementary interpretation is that the effects of mental practice on compensatory cognitive-motor mechanisms may develop more slowly than the immediate effects of physical practice. mental practice engages premotor, supplementary motor, parietal, prefrontal, and associative networks involved in motor planning, attention, visuospatial processing, and top-down control [14]. These regions overlap with networks recruited by people with PD to compensate for impaired automatic gait control [7,11]. Repeated mental rehearsal combined with physical execution may therefore have gradually strengthened the functional use of these compensatory networks, with measurable behavioral effects becoming more evident only at follow-up. Under this interpretation, the absence of immediate superiority does not necessarily indicate absence of effect, but may suggest that the contribution of mental practice is more closely related to retention, consolidation, or delayed stabilization of cognitive-motor strategies.

The improvement in global cognitive performance observed in the experimental group provides additional support for this interpretation. Although the T-MoCA is a global screening measure and does not allow domain-specific conclusions, the result suggests that the intervention including mental practice may have engaged cognitive processes beyond those required for physical execution alone. Mental practice requires sustained attention, internal simulation, working memory, visuospatial processing, anticipation of task demands, and strategic planning of movement. These processes are closely related to executive functions, which play a critical role in gait regulation in people with PD, particularly when automatic gait control is impaired and walking becomes increasingly dependent on goal-directed control. The sustained improvement in T-MoCA scores may therefore have contributed to the maintenance of benefits observed in Rapid Turns Test performance and perceived mobility, although this mechanism should be confirmed in future studies using domain-specific cognitive and cognitive-motor measures.

Another relevant aspect of the findings is the dissociation between broad self-reported clinical measures and outcomes more directly related to cognitive-motor performance and mobility. Self-reported FOG severity and motor aspects of daily living improved over time without differential effects between groups, suggesting that both interventions were capable of producing perceived benefits. This is plausible because both groups received structured FOG education, repeated therapist contact, remote supervision, and the same physical practice protocol. However, the absence of between-group differences in NFOG-Q and MDS-UPDRS Part II does not contradict the differential effects observed in Rapid Turns Test performance, T-MoCA, and the PDQ-39 mobility domain. Rather, it suggests that the added value of mental practice may have been more specific to context-dependent gait regulation, internal strategy use, and maintenance of performance under challenging conditions. Performance-based measures, such as the Rapid Turns Test, may capture aspects of FOG behavior that are not fully reflected in questionnaires, particularly because FOG is episodic, context-dependent, and influenced by retrospective recall.

From a mechanistic perspective, Dynamic Neuro-Cognitive Imagery may have shaped the mental practice component by increasing the multisensory, kinesthetic, and context-specific nature of motor imagery. Rather than simply adding more practice time, Dynamic Neuro-Cognitive Imagery may have helped participants construct more accessible and functionally relevant internal representations of movement, particularly in situations resembling common FOG triggers. This interpretation is consistent with previous studies suggesting that Dynamic Neuro-Cognitive Imagery can influence imagery ability, motor performance, cognition, and body representation in people with PD and in non-clinical populations [16,19,20]. In the present study, this may help explain why the added value of mental practice was observed mainly in the maintenance of gains and in outcomes requiring cognitive-motor integration, rather than as broad immediate superiority across all measures.

The remote delivery format is also clinically relevant. Telerehabilitation allowed the intervention to be delivered in participants’ home environments, where many FOG triggers occur and where strategies need to be applied in daily life. This context may have supported the transfer of learned strategies to real-world mobility situations. At the same time, remote interventions depend on technological access, participant engagement, environmental safety, and, in some cases, caregiver support. Therefore, although the present findings support the potential of remotely delivered cognitive-motor rehabilitation for people with PD and FOG, implementation in routine care should consider digital literacy, supervision needs, and individual clinical profiles.

Overall, these findings suggest that improving gait performance during supervised practice may not be sufficient for sustained FOG management. Interventions that combine task-specific physical training with strategies targeting motor imagery, cognitive control, and context-specific adaptability may be more likely to support retention and maintenance of functional gains. In this sense, mental practice grounded in Dynamic Neuro-Cognitive Imagery principles may represent a promising approach to complement physical practice in people with PD and FOG.

### Strengths and Limitations

This study has important methodological strengths. It used a randomized controlled design to investigate a mechanism-driven intervention combining mental practice, grounded in Dynamic Neuro-Cognitive Imagery principles, with physical practice in people with PD and FOG. A key strength was that both groups received the same therapist contact, remote supervision, FOG education, and structured physical practice protocol. Thus, the main planned difference between groups was the replacement of seated upper-limb stretching in the control group by mental practice/Dynamic Neuro-Cognitive Imagery in the experimental group. Because the stretching component was not designed to target gait or FOG mechanisms, this design strengthens the interpretation that the differential effects observed in the experimental group were related to the added cognitive-motor component rather than to differences in training time, attention, or exposure to physical practice. The study also included standardized assessments at baseline, immediately after intervention, and follow-up, allowing examination of both immediate and short-term retention effects.

This study also has limitations. All assessments and interventions were performed remotely, without an in-person physical examination, which limited direct assessment of motor signs, postural control, gait quality, and safety under standardized clinical or laboratory conditions. The study did not include neurophysiological measures, such as fNIRS, EEG, or functional neuroimaging, or detailed kinematic/kinetic assessments; therefore, the proposed cognitive-motor mechanisms should be interpreted as plausible explanations rather than demonstrated mechanisms. The follow-up period was limited to 30 days, preventing conclusions about long-term retention. Finally, because participation required internet access, video-call capability, and adequate home safety conditions, the findings may not generalize to people with more advanced PD, severe FOG, substantial cognitive impairment, limited digital literacy, or unsafe home environments.

### Clinical implications

The findings suggest that adding mental practice grounded in Dynamic Neuro-Cognitive Imagery principles to physical practice may be clinically useful for supporting the maintenance of gains in people with PD and FOG. Rather than replacing physical training, mental practice may complement physical practice by helping participants rehearse internally generated strategies to anticipate FOG triggers, organize movement, and apply compensatory responses beyond supervised sessions. This may be particularly relevant because maintaining improvements after the end of rehabilitation remains a major challenge in FOG management.

The remote delivery format also has important clinical implications. A structured mental practice/Dynamic Neuro-Cognitive Imagery plus physical practice protocol can be delivered synchronously at home with minimal equipment, potentially expanding access to specialized rehabilitation for people with mobility, transportation, geographic, or financial barriers. However, remote implementation should include safety procedures, therapist supervision, attention to digital literacy, and individualized assessment of the home environment. Future studies should refine intervention dose, identify which cognitive and motor profiles benefit most, and test whether hybrid models can further improve long-term adherence and clinical effectiveness.

## Conclusion

This study suggests that adding mental practice grounded in Dynamic Neuro-Cognitive Imagery principles to physical practice may support more stable short-term gains in people with PD and FOG. Both groups showed improvements after training, but the combined intervention was associated with better maintenance of performance during a provocative turning task, as well as favorable changes in global cognitive performance and mobility-related quality of life. These findings indicate that mental practice/Dynamic Neuro-Cognitive Imagery may be a useful adjunct to physical practice, particularly for interventions aimed at reinforcing internal strategies for FOG management. Further studies with longer follow-up periods, larger and more diverse samples, and neurophysiological or instrumented gait measures are needed to confirm these findings and clarify the mechanisms underlying the observed effects.

## Author Contributions

MEPP, PRS, and KYTH designed the study. MEPP, PRS, KYTH and LBRS designed the assessment protocol. PRS, LBRS, JMG and BHTS collected the data and were involved in data processing and data analysis. Statistical analysis was performed by MEPP. The draft of the manuscript was prepared by PRS and LMA, and was critically reviewed by MEPP. All authors confirmed and approved the final version of the manuscript.

## Data Availability

The datasets generated, used and analyzed during the trial are or will be available from the corresponding author upon reasonable request.

## Acknowledgments

This paper is dedicated to the memory of Professor Antonio Galves, whose unwavering commitment to the CEPID project laid the foundation for a multidisciplinary research group destined to thrive for years to come.

The study was supported by São Paulo Research Foundation (FAPESP) through the Research, Innovation and Dissemination Center for Neuromathematics (CEPID NeuroMat, grant number 2013/07699-0 and individual support by FAPESP for the author PRS by grant number 2025/14403-7.

The authors sincerely thank all participants for their collaboration and commitment, as well as the technical and administrative teams who provided essential support in carrying out the research activities. We also acknowledge the institutions involved for their continuous support in the development of this work.

## Notes

### Competing Interest Statement

The authors have declared no competing interest.

### Clinical Trial

NCT06957405

### Clinical Protocols

https://www.medrxiv.org/content/10.1101/2025.04.28.25326452v1.article-metrics

### Funding Statement

The study was supported by Sao Paulo Research Foundation (FAPESP) through the Research, Innovation and Dissemination Center for Neuromathematics (CEPID NeuroMat, grant number 2013/07699-0 and individual support by FAPESP for the author PRS by grant number 2025/14403-7.

### Author Declarations

The study was approved by the Research Ethics Committee of the School of Medicine of the University of Sao Paulo under protocol number 7.253.458.

